# A loss-of-function mutation in the GTPase domain of *MFN2*, perverting mitochondrial dynamics, is associated with dilated cardiomyopathy

**DOI:** 10.64898/2026.08.10.26360061

**Authors:** Mohini Gupta, Amrita Mukhopadhyay, Manohar Lal Yadav, Dharmendra Jain, Bhagyalaxmi Mohapatra

**Affiliations:** Cytogenetics Laboratory, Department of Zoology, Institute of Science, Banaras Hindu University, Varanasi-221005, Uttar Pradesh, India; Department of Cardiology, Institute of Medical Sciences, Banaras Hindu University, Varanasi-221005, Uttar Pradesh, India

**Author notes:** Corresponding Author: Bhagyalaxmi Mohapatra Email Address).

## Abstract

Mitofusin 2 (MFN2), a key outer mitochondrial membrane GTPase, regulates mitochondrial fusion, mitophagy, calcium homeostasis, and cellular bioenergetics. This study investigated the role of *MFN2* variants in patients with Dilated Cardiomyopathy (DCM) using whole-exome sequencing (WES) of 5 familial and 10 sporadic DCM cases.

A rare de-novo *MFN2* variant, c.932A>G (p. N311S), was identified in a DCM patient, which is absent in 100 healthy controls as well as in the 1000 Genomes, IndiGenomes, databases while it shows very low MAF (0.0000081) in gnomAD. Structural modelling predicted the variant to be highly deleterious and revealed marked conformational distortion of the mutant protein (RMSD = 8.95 Å). Molecular docking further showed a weakened interaction between MFN2-N311S and PRKN (Parkin), indicating impaired mitophagy and defective mitochondrial quality control. Moreover, functional analysis in stable H9c2 cardiomyoblast cell lines demonstrated significantly reduced MFN2 mutant protein expression, extensive mitochondrial clustering and fragmentation. The mutant protein also indicated significant reduction in mitochondrial membrane potential, ATP production, and oxygen consumption rate (OCR), together with elevated cytosolic Ca² and reactive oxygen species (ROS) levels. qRT-PCR analysis further revealed activation of the PI3K/AKT/mTOR signalling pathway and increased expression of hypertrophic markers *Myh6, Nppa, Nfatc1*, and *Nfatc2*.

The above findings collectively highlight the significant impact of the *MFN2* mutation on mitochondrial dynamics and cellular health, suggesting a significant correlation with the pathogenesis of DCM. This finding could further open a door to develop a potential therapeutic target for DCM.

## 1. Introduction

Dilated cardiomyopathy (DCM) is a myocardial disorder characterized by left ventricular chamber enlargement and impaired systolic function, depicted as a leading cause of heart failure with a prevalence of 1 in 2,500 individuals (Codd et al., 1989; Taylor et al., 2006; Jefferies & Towbin, 2010) of which approximately 30–40% are familial, most often inherited in an autosomal dominant manner (Keeling et al., 1995; Giri et al., 2022).To date, many pathogenic variants have been identified in a wide spectrum of genes encoding components of the sarcomere, cytoskeleton, nuclear envelope, ion channels, sarcoplasmic reticulum, and desmosomal complexes. However, in recent years, increasing attention has been directed toward the contribution of mitochondrial dysfunction and mutations in mitochondrial genes as emerging drivers of DCM pathogenesis, opening novel insights for mechanistic and therapeutic interventions.

Mitochondria supply the energy in the form of ATP to cardiac muscles to pump blood efficiently. In order to meet the energy supply and demands of cardiac muscle, mitochondria regulate their fusion and fission processes. Fusion of mitochondria is known to be mediated by Mitofusins namely, Mitofusin1(MFN1) and Mitofusin2 (MFN2) on the outer mitochondrial membrane (OMM) along with another fusion-protein Optic atrophy 1 protein (OPA1) on the inner mitochondrial membrane (IMM) (Song et al., 2009; Hall et al., 2014; Tilokani et al., 2018). Among these, MFN2 is a dynamin-related GTPase that plays a pivotal role in maintaining mitochondrial homeostasis, orchestrating essential processes such as oxidative metabolism, ER-mitochondrial tethering, mitophagy, axonal trafficking, and cell cycle regulation (Ngoh et al., 2012; Chen & Dorn, 2013; Chen et al., 2014; Schrepfer & Scorrano, 2016; Chandhok et al., 2018; Green et al., 2022). Structurally, the MFN2 protein consist of 757 amino acid that comprises four distinct domains: a highly conserved N-terminal GTPase domain (aa 93-346) responsible for nucleotide binding and hydrolysis; and two coiled-coil domain [Heptad Repeats (HR)) namely, HR1(aa 391-434) and HR2(aa 695-738)] facilitating membrane tethering and fusion, and a C-terminal with a bipartite transmembrane domain which span between aa 605-647 that anchor the protein to the outer mitochondrial membrane (OMM). Additionally, the protein has an N-terminal p21ras signature motif (aa 77-96) and a proline-rich (PR) domain located between HR1 and the TM regions is implicated in mediating protein–protein interactions, underscoring multifunctional regulatory capacity of MFN2 (Detmer & Chan, 2007; Qi et al., 2016; Chandhok et al., 2018; Filadi et al., 2018; Lv et al., 2025).

MFN2 mutations were first identified in Charcot–Marie–Tooth disease type 2A (CMT2A) and are also associated with Alzheimer’s disease, Parkinson’s disease, several cancers, and cardiovascular disorders. (Züchner et al., 2004; Wang et al., 2009; Chen et al., 2011; Zhang et al., 2013; Li et al., 2018; Lv et al., 2025). Experimental models have demonstrated that cardiac-specific, conditional ablation of *Mfn1* and *Mfn2* in adult mice leads to lethal dilated cardiomyopathy, underscoring their indispensable role in maintaining myocardial integrity (Papanicolaou et al., 2012). Notably, *Mfn2* deficiency alone in adult murine hearts precipitates DCM accompanied by impaired mitophagy, highlighting its essential function in mitochondrial turnover and cardiac homeostasis (Chen et al., 2011).

In this study, we have investigated the potential role of *MFN2* in DCM. Through whole-exome sequencing of DCM patients, we identified a rare *MFN2* variant (p.N311S) and evaluated its pathogenicity using integrated *in-silico* and *in-vitro* approaches. Structural modelling predicted significant alterations in both the secondary and tertiary structures of the mutant protein. Functional analyses demonstrated that the p.N311S variant resulted in impaired mitochondrial membrane potential, reduced ATP production, decreased oxygen consumption rate (OCR), elevated cytosolic calcium and reactive oxygen species (ROS) levels, and enhanced mitochondrial fragmentation compared with the wild-type (WT) protein. Transmission electron microscopy (TEM) further revealed abnormal perinuclear accumulation of mitochondria in cells expressing the mutant MFN2. Collectively, these findings indicate that the *MFN2*-N311S variant disrupts mitochondrial homeostasis by impairing mitochondrial bioenergetics, dynamics, and intracellular distribution, ultimately contributing to cardiac dysfunction. To the best of our knowledge, this is the first study to identify and functionally characterize an *MFN2* variant associated with DCM in the Indian population.

## Material & Methods

### 2.1 Clinical Evaluation and Enrolment of Patients

Five multigenerational families, and 10 sporadic cases as well as 100 ethnic-matched healthy control individuals (without any cardiac disease) were recruited from the Department of Paediatric Medicine and Department of Cardiology, Institute of Medical Sciences, Banaras Hindu University. The diagnosis of DCM was established using a combination of inclusion and exclusion criteria, with primary emphasis on left ventricular (LV) size and function, specifically, a fractional shortening (FS) of less than 25% and an ejection fraction (EF) below 45%. All study subjects were evaluated by physical examination, i.e. two-dimensional (2D) echocardiography, electrocardiogram (ECG), and cardiac magnetic resonance imaging (cardiac MRI). The study was approved by the Institutional Ethics Committee.

### 1.2 Whole exome sequencing (WES)

Trio-based WES was conducted, involving the probands and their first-degree relatives. Genomic DNA was extracted from the collected blood sample, and quality of DNA was checked and estimated in a Nanonodrop 2000 (Thermofisher Scientific). For whole exome sequencing, proband along with at least one affected and one unaffected member (internal control) were selected. Library preparation for WES was performed using the ‘Human Nextera Rapid Capture Expanded Exome Kit’ (Illumina, USA), in accordance with the manufacturer’s guidelines. The prepared libraries were sequenced using the Illumina HiSeq 2500 platform (Illumina, USA), generating 2X150 bp paired-end reads per sample, with an average coverage depth of 100X.

### 2.3 NGS data analysis and variant prioritization

Data analysis commenced upon completion of sequencing. Sequencing reads were aligned to the human reference genome (GRCh38: Genome Reference Consortium Human Build 38). Subsequent realignment around known insertion-deletion sites was carried out using the Genome Analysis Toolkit (GATK), and base quality score recalibration was performed using the GATK recalibrator. Variant Call Format (VCF) files were then generated for each individual sample. A detailed description of the NGS data analysis workflow is available upon request. The potential deleterious variants were prioritized using the pipeline. Briefly, the variant with read-depth ≥ 20 were selected to avoid the loss of potential variants with low read depth. An inhouse curated cardiac gene panel, including genes involved in mitochondrial form and function, was applied to select the variants. The variants with minor allele frequency (MAF) ≥ 0.01 in the Genome Aggregation Database (gnomAD; https://gnomad.broadinstitute.org/), 1000 Genome allele frequency Projects (https://www.internationalgenome.org/), Exome Aggregation Consortium (ExAC) allele frequencies database (http://exac.broadinstitute.org/), IndiGenomes database (https://clingen.igib.res.in/indigen/), INDEX database (https://indexdb.ncbs.res.in/search/) were removed. Further selected variants were categorized into missense, stop codon, frameshift, splice-site, synonymous, and intronic variants, depending on their effect on amino acid sequence.

### 2.3 *In-silico* analysis

#### 2.3.1 Prediction of pathogenic potential

The disease-causing potential of non-synonymous variant identified in *MFN2* was predicted using VarCards (http://www.genemed.tech/varcards/search), which incorporates more than 20 *in-silico* predictive algorithms e.g., PolyPhen-2 (http://genetics.bwh.harvard.edu/pph2/), PANTHER (http://www.pantherdb.org/tools/csnpScoreForm.jsp), SIFT (http://sift.bii.astar.edu.sg/www/SIFT_seq_submit2.html), Mutation Taster (http://www.mutationtaster.org), I-Mutant (http://folding.biofold.org/i-mutant/i-mutant2.0.html), FATHMM (http://fathmm.biocompute.org.uk/fathmmxf/index.html), VEST3 (https://karchinlab.org/apps/appVest.html) and M-CAP (http://bejerano.stanford.edu/mcap/), PROVEAN (Protein Variant Effect Analyzer, http://provean.jcvi.org/index.php), CADD (https://cadd.gs.washington.edu/), and SNAP2 Screening of Non Acceptable Polymorphism, (https://rostlab.org/services/snap2web/) generates a combined damaging score from 0-1, called Varcards score. The variant with a damaging score of 1 are highly damaging, while those with a score 0 are non-damaging.

#### 2.3.2 Phylogenetic conservation of amino acid residues in MFN2 protein

The reference genomic DNA sequence (NC_000001.11), mRNA sequence (NM_014874.4) and protein (NP_001121132.1) sequence of MFN2 were listed from Gen-Bank (https://www.ncbi.nlm.nih.gov/genbank/) and Protein database (https://www.ncbi.nlm.nih.gov/protein/) for all the *in-silico* analyses. Multiple sequence alignment of MFN2 protein (NP_001121132.1) homologs across different species was performed using the ‘HomoloGene’ feature of NCBI to evaluate the conservation of substituted amino acids.

#### 2.3.3 Secondary structure prediction and three dimensional (3-D) modelling

Secondary and tertiary structure of a protein depends on the sequences and properties of its constituent amino acid residues. Non-synonymous changes affect the structures of proteins depending upon the level of differences in their side chains. Therefore, the secondary structural conformation of WT and mutated MFN2 protein was predicted by PsiPred (http://bioinf.cs.ucl.ac.uk/psipred/) bioinformatics tool. Simultaneously, three-dimensional homology modelling of WT and MFN2 mutant protein was performed by iTASSER to obtain their suitable structure. Both, WT and mutated models were generated with a confidence level of greater than 90%. Further, modelled structures were aligned with the help of UCSF Chimera and RMSD value was calculated.

#### 2.3.4 Protein-protein docking

To elucidate the molecular interaction between MFN2 and PRKN (Parkin), molecular docking was conducted using the HADDOCK2.4 web server (Dominguez et al., 2003). The PDB structures of both interacting partners were uploaded to the server, and active (binding) residues were defined based on previously reported literature (Chen & Dorn, 2013; Gong et al., 2015; Song et al., 2015; Franco et al., 2023) . The resulting docked complexes were subsequently analyzed and visually inspected using PyMOL (version 4.6) to assess interfacial contacts, and structural perturbations induced by the mutations.

#### 2.3.5 Prediction of putative phosphorylation site

Phosphorylation of threonine, tyrosine, and serine is one of the most important and ubiquitous post-translational modifications of proteins that regulate several biological processes such as signal transduction, cell-cycle and proteolysis etc. Therefore, potential phosphorylation sites in both WT and MUT protein were predicted by NetPhos3.1 Server (http://www.cbs.dtu.dk/services/NetPhos-3.1/). The NetPhos 3.1 server predicts serine, threonine or tyrosine phosphorylation sites for 17 kinases (ATM, CKI, CKII, CaM-II, DNAPK, EGFR, GSK3, INSR, PKA, PKB, PKC, PKG, RSK, SRC, cdc2, cdk5 and p38MAPK) in eukaryotic proteins using ‘Ensembles of neural networks’. Prediction score values range from 0 to 1.0 and amino acids having a prediction score above 0.5 were considered as potential phosphorylation sites.

### 2.4 *In-vitro* characterization of the N311S variant of MFN2

#### 2.4.1 Cloning of wild type *MFN2* and Site Directed Mutagenesis

MFN2 full length clone was a kind gift from Prof. Pozzan’s Lab (University of Padova, Italy), which was sub-cloned in the mammalian expression vector pcDNA3.1/NT-GFP-TOPO™ (Thermofisher,) vector, following the manufacturer’s protocol. The fidelity of the MFN2 construct was confirmed through Sanger sequencing. WT-MFN2 was used as template for preparing mutant constructs of MUT-MFN2 by site-directed mutagenesis using Quick change II XL site-directed mutagenesis kit (Agilent Technologies Inc., Santa Clara, CA, USA) as per manufacturer’s instructions. Introduced mutation was confirmed by Sanger sequencing.

#### 2.4.2 Transfections and generation of stable cell lines

H9c2, embryonic rat heart-derived myoblasts, was cultured in Dulbecco’s modified Eagle’s medium (DMEM Gibco, Life technologies corp., NY, USA), enriched with 10% fetal bovine serum (FBS Gibco, Life technologies corp., NY, USA), in a 95% air and 5% CO atmosphere. To establish stable cell lines expressing both wild type and mutant *MFN2* constructs, we started with a kill-curve experiment to identify the optimal concentration of G418 for selection. After seeding the cells, transfection of constructs with the WT and MUT (N311S) plasmids was accomplished using Fugene 6 transfection reagent (Promega, Corporation) as per the user manual. G418 was added to the culture to maintain selection pressure. Over time, distinct colonies emerged, indicating successful integration of the genes. These healthy colonies were isolated and cultured in G418-supplemented media, ensuring stable expression of the MFN2 constructs, also confirmed by their positive GFP expression.

#### 2.4.3 MitoTracker staining

In order to perform immunostaining for visualizing cellular localization and mitochondria assembly of MFN2-WT and MFN2-MUT stable cells. These cells were seeded on glass coverslips inside six well culture plates and grown in Dulbecco’s modified Eagle’s medium (DMEM, Gibco, Life technologies corp., NY, USA), supplemented with 10% Fetal Bovine Serum (FBS, Gibco, Life technologies corp., NY, USA), incubated in 37 C at 5% CO2, overnight. Next, cells were washed with 1X PBS twice and incubated with 50 nM MitoTracker Red CMXROS (Molecular Probes) for 30 minutes at 37 C and then fixed with 4% paraformaldehyde (PFA) for 15 minutes, followed by permeabilization with 0.5% Triton X (diluted in 1X PBS). After blocking with 5% milk (in 1X PBS) for 2 hours, cells were washed with 1X PBS 5-6 times, followed by nuclei were stained with DAPI, mounted with mounting media and imaging was carried out using Zeiss LSM 510 Meta, Laser Scanning Confocal microscope, further analysed with utilizing ImageJ to analyze the index of elongation.

#### 2.4.4 Transmission electron microscopy

Stable cell lines of MFN2 constructs WT and MUT were grown and fixed in 2.5% glutaraldehyde and 2% paraformaldehyde in 0.1M sodium phosphate buffer (pH 7.4) initially for 20 minutes at room temperature and then for 4-6 hours at 4^0^C. The samples were dehydrated in acetone, infiltrated in toluene and resin and finally embedded in Araldite CY212 r e s i n (TAAB, UK). Thin sectioned (60-70 nm) stained with 1% aqueous uranyl acetate and 0.5% alkaline lead citrate (pH 13) for 10 minutes in each step, washed gently with distilled water and mounted onto 300 mesh copper grids. The grids were observed under a TALOS F200S transmission electron microscope (Thermo Fisher Scientific, Waltham, MA, USA) at an operating voltage 200 kV. Images were digitally acquired at suitable magnifications (2050X to 5500X) using TIA software attached to the microscope. Mitochondrial size and surface density were quantified using ImageJ (NIH)

#### 2.4.5 Western Blotting

With a view to estimate the expression level of MFN2 protein in wild type versus mutant, stably transfected H9c2 cells were seeded in six well plate. After 24 hr, cell-lysates were prepared in RIPA buffer (50 mM Tris-HCl, pH 8.0, 150 mM NaCl, 0.1% Triton X, 0.5% sodium deoxycholate, 0.1 % SDS) with 1mM sodium orthovanadate, 1 mM NaF, and protease inhibitor tablets (Roche, Basel, Switzerland). After dilution with Laemmeli’s sample buffer, the lysates were separated on 10 % polyacrylamide SDS gel and proteins were transferred to PVDF membrane (Bio-Rad Laboratories Inc, CA, US). The PVDF membrane was blocked for 2 hours in 5 % dry skimmed milk at room temperature, followed by overnight incubation with anti-MFN2 antibody (CST-Aldrich, MO, US) at 4 C, the blot was washed with PBST (0.1 % Tween-20 diluted in 1X PBS) six times for 10 mins each. The blot was then incubated with HRP conjugated goat anti-rabbit IgG antibody (Genei, Merck Specialties Pvt. Ltd., Darmstadt, Germany) followed by washing with PBST (6X, 5 min each) and detected with ECL detection kit (GE Healthcare, IL, US).

#### 2.4.6 MTT assay

Stable cell lines of MFN2-WT and MFN2-MUT were seeded in 96 wells plate at density of (∼2 X 10^4^). After 24 hrs 0.5 mg/ml MTT was added in each well, incubated at 37°C in 5% CO_2_ for 4-5 hrs. The blue formazan was dissolved in DMSO and incubated in dark for 10 mins. Formazan accumulation exposes mitochondrial activity, which indirectly affects cell’s ability to survive (Riss et al., 2016). The final purple color was assessed by measuring absorbance at 570 nm in microplate reader (Bio-Rad) and cell viability was calculated using formula:

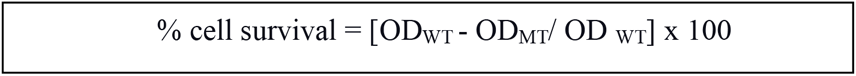

OD (Optical Density) is the absorbance value measured by the spectrophotometer, which reflects the amount of purple formazan produced and thus the number of viable cells in the MTT assay.

#### 2.4.7 Mitochondrial membrane potential

Stable cell lines of MFN2-WT and MFN2-MUT were grown in 6 well plate. After 24 hours, respective cells were washed with PBS, trypsinized and collected in 1.5 ml micro-centrifuge tubes and then stained with 200 nM TMRE dye (Tetramethylrhodamine ethyl ester) (Invitrogen #Catalog number T669) in 1XPBS. The sample were then incubated at 37°C in dark for 30 min. Flow cytometry FACS (CytoFLEX LX Bachman Counter) reading was taken for the cells, stained with orange fluorescent dye and intensity was measured which correspond to the membrane potential of the stained cells. The data are expressed as a geometric mean fluorescence intensity (gMFI) ± SD from(n=6) independent experiments.

#### 2.4.8 ATP content

WT and MUT stable cell lines of *MFN2* were grown in 12 well plates in equal numbers (10^2.5^ cells) for 24 hours. Cells were washed with 1X PBS twice, and ATP content was measured using the ENLIGHTEN ATP assay system according to the manufacturer’s instructions (Promega, WI, USA). Briefly, we extracted ATP from the cells using 0.5% TCA and neutralized it with 0.1 M Tris-acetate buffer pH 7.75. After adding an equal volume of rL/L recombinant luciferase reagent (ENLITEN® ATP Assay System, Promega), luminescence readings were taken by using an illuminometer with a delay of 2 sec and 10 sec as a readout for ATP levels. The data are expressed as a geometric mean fluorescence intensity (gMFI) ± SD from (n=6) independent experiments.

#### 2.4.9 Mitochondrial mass with NAO

Stable cell lines of MFN2-WT or MFN2-MUT were grown in 6 well plates containing equal numbers of cells (10^5^/ well) for 24 hours. Cells were trypsinized and suspended in the fresh culture medium supplemented with 1 μM NAO (Invitrogen), incubated at 37°C in 5% CO2 for 30 minutes, according to (Maftah et al., 1989). After washing with 1XPBS, the cells were suspended in PBS with calcium and magnesium ions. Measurements were done in FACS CAlibur with excitation/emission 495/522 nm. For each sample 10,000 events were counted. The data are expressed as a geometric mean fluorescence intensity (gMFI) ± SD from (n=6) independent experiments.

#### 2.4.10 Flow cytometry of total ROS

MFN2 stable cell lines of WT and MUT were grown in 6 well plates (10^5^ cells/well) for 24 hours. Cells were trypsinized and suspended in the fresh culture medium supplemented with MitoSOX Green (5 μM for 10 min at 37°C). Fluorescence was analyzed on a ‘FACS CAlibur Flow Cytometer’ (Bio-Rad). Data were analyzed as geometric mean and are presented as mean fluorescence intensity of (n=6) independent experiments.

#### 2.4.11 Quantitative Analysis of Mitochondrial Oxygen Consumption Rate (OCR)

Mitochondrial respiration was measured using a high-resolution respirometer (Oxygraph-2 k, OroborosInstruments) at 37°C under stirring conditions (750 rpm). MFN2 stable cell lines of WT and MUT cells were trypsinized, washed twice and resuspended mitochondria respiration buffer. A total of 1X10^6^ cells were introduced into each oxygraph chamber, with separate chamber containing MFN2-WT and MFN2-MUT. Respiration was allowed to determine routine respiration, reflecting the physiological coupling state under endogenous substrate supply and ATP demand. Then, Cells were then sequentially treated with oligomycin (1 μg/ml), FCCP (0.5 µM), and antimycin A (1 μg/ml) to study leak respiration, maximum capacity of electron transport system and residual/extra-mitochondrial oxygen consumption, respectively. Calibration at air saturation was performed each day before starting experiments by letting Buffer B stir with air in the oxygraph chamber until equilibration and a stable signal was obtained. All experiments were performed at an oxygen concentration in the range of 100–205 μM O_2_. Data were recorded and analyzed using DatLab 7.4 software (Oroboros Instruments).

#### 2.4.12 Cytoplasmic Ca^+2^ measurement

Stable cell lines for MFN2-WT and MFN2-MUT were grown in 6 well plates (as described above) for 24 hours. Cells were trypsinized and suspended in the fresh culture medium supplemented with Fluro 2AM (Invitogen Catalog number F1201) (0.5 μM for 30 min at 37°C). Fluorescence was analyzed on a ‘FACS CAlibur Flow Cytometer’ (Bio-Rad). Data were analyzed as geometric mean and are presented as mean fluorescence intensity of (n=6) independent experiments.

#### 2.4.13 Quantative PCR

For real time PCR assay, both MFN2-WT and MFN2-MUT, stably transfected cells were grown in 6 well plate in equal number (1×10^6^ cells well) for 24 hrs (Ambion, Thermo Fisher Scientific, USA) according to the manufacturer’s instructions. The isolated RNA was treated with RNase-free DNase I (ThermoFisher Scientific, USA), to remove genomic DNA contamination. RNA concentration and purity were determined using a NanoDrop™ 2000 spectrophotometer (Thermo Fisher Scientific, USA). First-strand cDNA was synthesized from 1 μg of DNase I-treated RNA using the RevertAid First Strand cDNA Synthesis Kit (Thermo Fisher Scientific, USA) according to the manufacturer’s protocol. Quantitative real-time PCR (qRT-PCR) was performed using the KAPA SYBR® FAST qPCR Master Mix Kit (Merck KGaA, Darmstadt, Germany) on a QuantStudio™ 5 Real-Time PCR System (Applied Biosystems, USA). Gene-specific primers were used to analyze the expression of genes involved in the PI3K/AKT signalling pathway and cardiac hypertrophic marker genes (*Myh6, Nappa, Actc1, Nfatc1 and Nfatc2*), with *GAPDH* serving as the endogenous reference gene for normalization. PCR specificity was determined using melting curve assessment and gene expression differences were determined using the 2−ΔΔCt method. All the experiments were performed in triplicates and expression data was plotted as fold change with the standard error of the mean. The significance of the data was calculated by unpaired Students’s *t*-test, with P < 0.05 considered statistically significant.

#### 2.4.14 Statistical Analyses

Data are expressed as the mean ± standard error of the mean (SEM). All the statistical test were performed and all graphs were constructed GraphPad Prism software 9.3.1. Statistical comparisons were conducted using unpaired t test. The level of significance was set to *P< 0.05, **P< 0.01 and ***P< 0.001.

## 3. Results

### 3.1 Identification of *MFN2* variant

Whole exome sequencing of 5 families (2 affected plus one unaffected individual per family) and 10 sporadic cases of DCM, revealed one missense variant in *MFN2* gene c.932A>G; (p. Asn311Ser,, rs748838916) with minor allele frequency (MAF 0.0000081). The proband was a 12 years old female with severely reduced LVEF=20% and increased LVIDd=88 mm, indicating extremely impaired left ventricular systolic function. The identified variant was further confirmed by bi-directional Sanger sequencing and segregation analysis was done in the family members. The variant was present only in proband, not in any other members, suggesting its *de novo* origin. Notably, mapping of this variant revealed its presence in the GTPase domain of MFN2 protein.

### 3.2 Phylogenetic conservation of *MFN2* across species

A cross species multiple sequence alignment of MFN2 protein viz., *Homo sapien* (NP_001121132.1), *Pan troglodytes*(XP_514395.3)*, Canis lupus familiarie* (XP_038513764.1), *Equus caballus* (XP_023491544.1), *Bos taurus* (NP_001019860.1), *Rattus norvegicus*(NP_001416898.1), *Mus musculus*(NP_573464.2) and *Gallus gallu* (XP_015152691.1) revealed that the substituted amino acid residue (Asparagine) was highly conserved. The conserved region with amino acid (N) along with flanking amino acid sequences has been shown in (Figure 2B).

**Figure 1.**
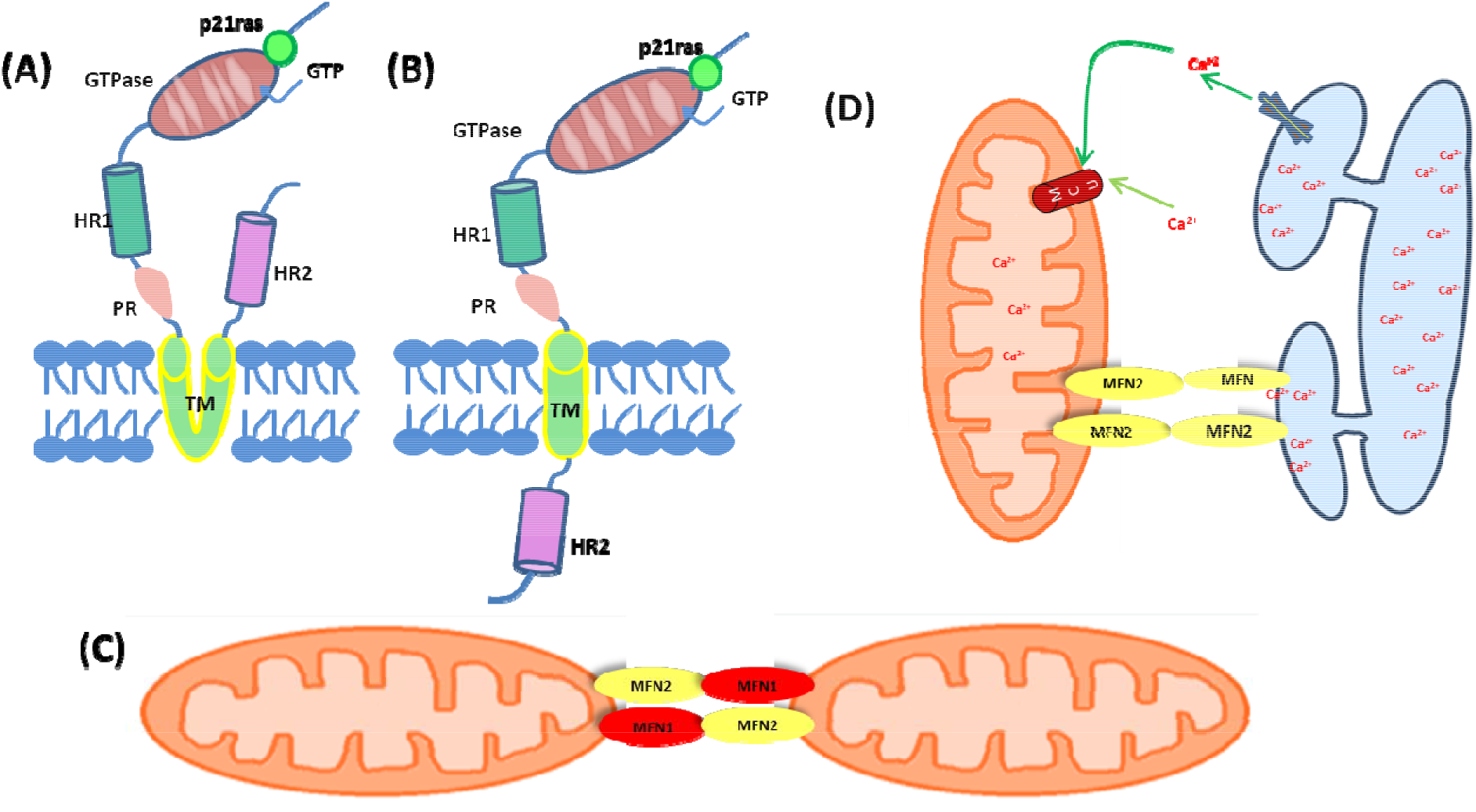
Structure and function of Mitofusin2 (MFN2) **(A)** Classical model of MFN2 topology showing the N-terminal GTPase domain, heptad repeat 1 (HR1), proline-rich (PR) domain, bipartite transmembrane (TM) domain, and C-terminal heptad repeat 2 (HR2) anchored in the outer mitochondrial membrane (OMM). **(B)** Revised topology model of MFN2, illustrating the proposed membrane orientation with the GTPase and HR1 domains exposed to the intermembrane space and the HR2 domain facing the cytosol. **(C)** MFN1 and MFN2 mediate mitochondrial outer membrane fusion through homo-oligomeric (MFN1–MFN1 or MFN2–MFN2) and hetero-oligomeric (MFN1– MFN2) interactions, promoting mitochondrial tethering and fusion in a GTP-dependent manner. **(D)** MFN2 mediates endoplasmic reticulum (ER)–mitochondria tethering at mitochondria-associated membranes (MAMs), facilitating Ca² transfer from the ER to mitochondria through the mitochondrial calcium uniporter (MCU) and thereby regulating mitochondrial calcium homeostasis and bioenergetics.

**Figure 2.**
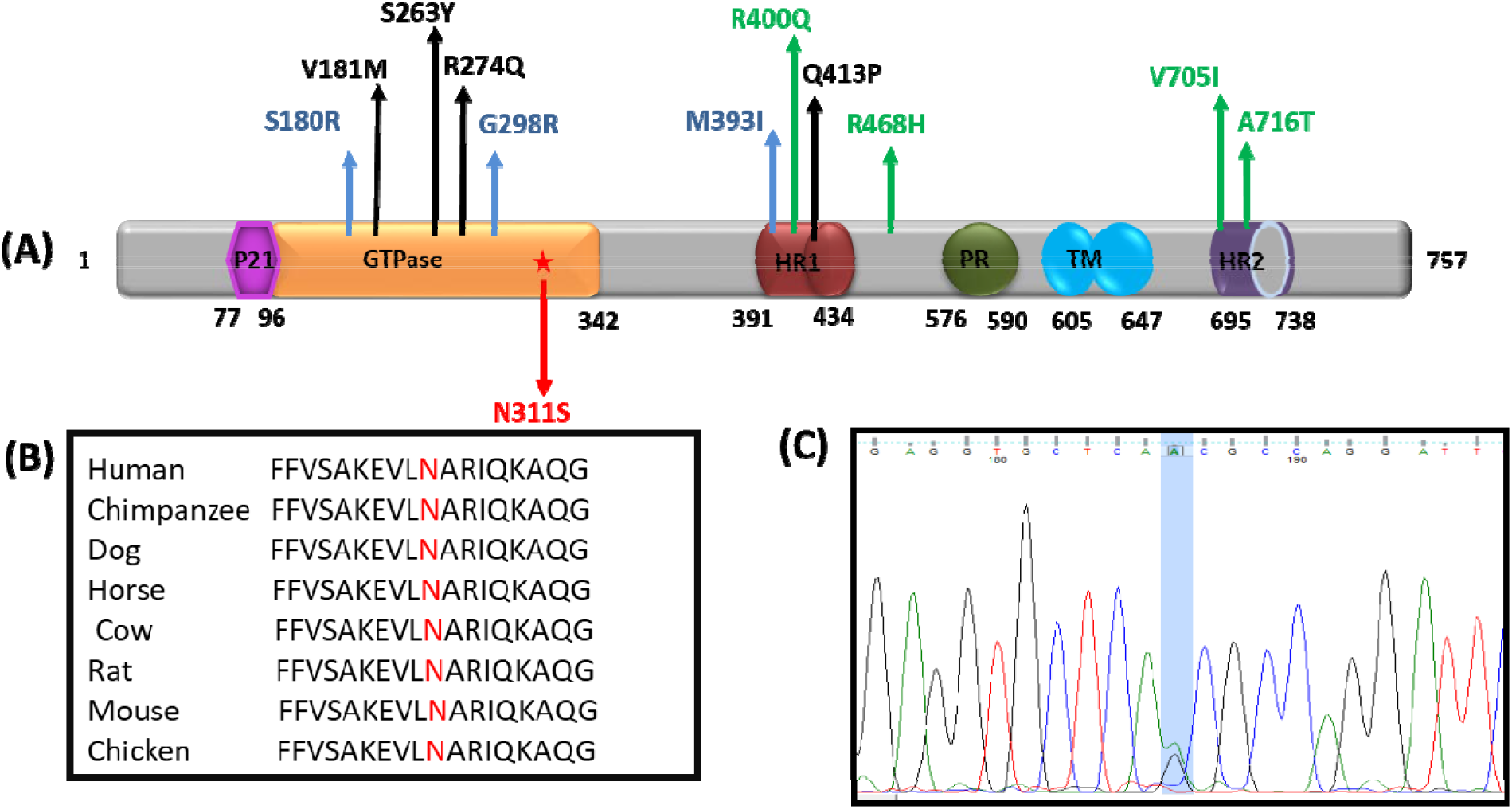
Schematic representation of MFN2 variant and their phylogenetic conservation **(A)** Diagrammatic representation of MFN2 protein (757 amino acid) showing different domains and modules of MFN2 protein, viz., p21ras (77-96 amino acids), GTPase (93-342 amino acids), HR1(Helical domain 1) (391-434 amino acids) domain, PR domain (576-590 amino acids) TM (transmembrane domain) (605-647 amino acids) and the HR2 (Helical domain 2) (695-738 amino acid). Position of identified non-synonymous are marked with red arrow. Previously reported MFN2 variants are color-coded based on their associated cardiac phenotype: blue for hypertrophic cardiomyopathy (HCM), black for dilated cardiomyopathy (DCM), and green for variants linked to both DCM and HCM. **(B)** Phylogenetic conservation of MFN2 protein across different species showing substituted amino acids (highlighted in red). **(C)** Sequence chromatogram of nonsynonymous variant c.932 CAA>CAG.

### 3.3 Predicted Conformational Alterations in MFN2 and Their Influence on PRKN Binding

The three-dimensional structures of both the wild-type (WT) and mutant (MUT) forms of the MFN2 protein were modelled and subsequently superimposed to assess the structural deviations induced by the mutation. Comparative structural analysis revealed a notable conformational change between the two models, as reflected by a root mean square deviation (RMSD) value of 8.951 Å (Figure 3A-C), indicative of substantial atomic displacement.

**Figure 3.**
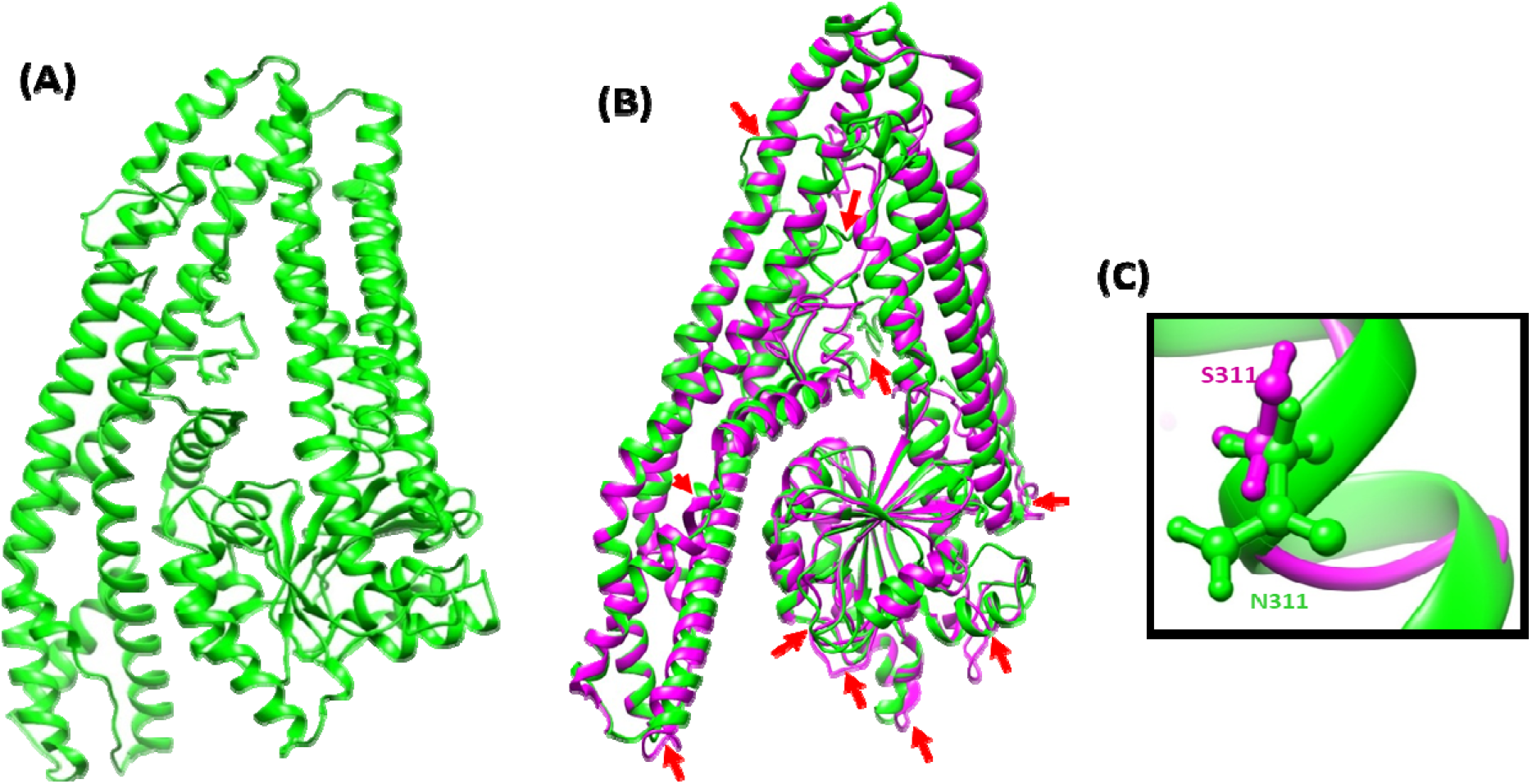
Structural comparison MFN2-WT and MFN2-MUT variant by UCSF Chimera. **(A)** Three-dimensional structures of MFN2-WT. **(B)** Superimposed three-dimensional structures of wild type (MFN2-WT) versus mutant protein (MFN2-MUT) using Chimera. The root mean square deviation (RMSD) value of the two superimpose structures is 8.951. WT tertiary structure of MFN2 protein has been shown in green color while MUT has bee shown in blue color. **(C)** WT and MUT three-dimensional structure showing amino acid, Asparagine (N) and MUT amino acid, Serine (S) with neighboring amino acids.

To examine the structural and functional implications of the N311S alteration on PRKN, we conducted *in silico* molecular docking simulations to assess the interaction between MFN2 and PRKN (Parkin), a crucial regulator of mitophagy. Molecular docking data indicated the MFN2-WT–PRKN complex exhibited an increased number of hydrogen bonds and a more cohesive interfacial interaction network, signifying enhanced binding affinity and superior structural stability. The N311S–PRKN complex demonstrated diminished hydrogen bonding and a compromised interaction contact.

The N311S mutation caused a spatial rearrangement of the binding interface. In the wild-type protein, PRKN interaction residues were primarily situated inside the N-terminal GTPase domain of MFN2, but in the mutant protein, the association transitioned to the C-terminal heptad repeat (HR) domains. The domain-level redistribution indicates that the N311S mutation may change the conformational landscape of MFN2, thus affecting its interaction dynamics with PRKN. Considering the documented function of the MFN2–PRKN connection in regulating mitophagy, such structural alterations may hinder mitochondrial quality control mechanisms and further intensify mitochondrial dysfunction. The detailed docking conformations and inter-domain interactions are illustrated in (Figure 4A-B).

**Figure 4.**
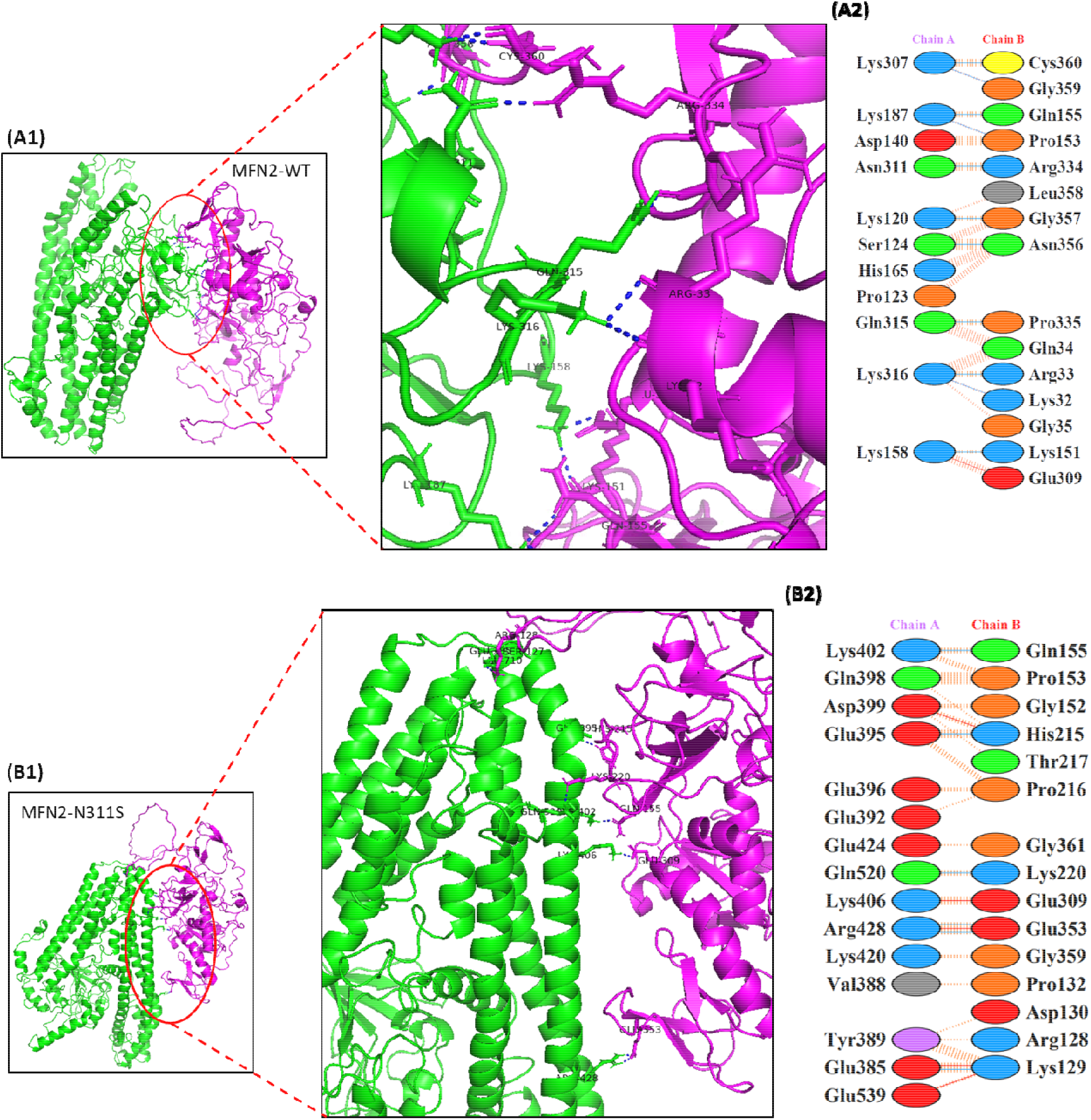
Protein-protein interaction of MFN2 protein with PRKN; We have shown protein-protein interactions of Wild-type MFN2 (Green) as well as mutant MFN2(N311S) with PRKN (Magenta). **(A1,B1)** represents protein-protein interaction, **(A2,B2)** represents residue involved in interaction.

### 3.4 Introduction of potential phosphorylation site at Ser311residue

Prediction of putative phosphorylation sites by NetPhos3.1 server has revealed that MFN2-wild-type protein possesses a total of 54 Serine residues of which 29 are considered as potential phosphorylation sites by different group of kinases. In p. Asn311Ser mutant protein, since aa residue, Asparagine, at 311^th^ position is substituted by Serine residue, total numbers of predicted phosphorylation sites have increased. The Ser311 was predicted to be phosphorylated by PKC and other unspecified kinases with prediction score of 0.616 and 0.871 respectively. The putative phosphorylation sites at Serine residues in MFN2 protein and specific kinases causing phosphorylation are given in (Figure 5). Each group of kinases requires specific amino acid sequences in order to perform their function. For example, PKC-kinases phosphorylate serine residues followed by arginine residues i.e. these kinases show arginine-directed phosphorylation activity. The p.N311S substitution modifies the wild-type sequence V-L-N-A-R-I-Q to V-L-S-A-R-I-Q which creates a potential phosphorylation site required by the PKC-kinases.

**Figure 5.**
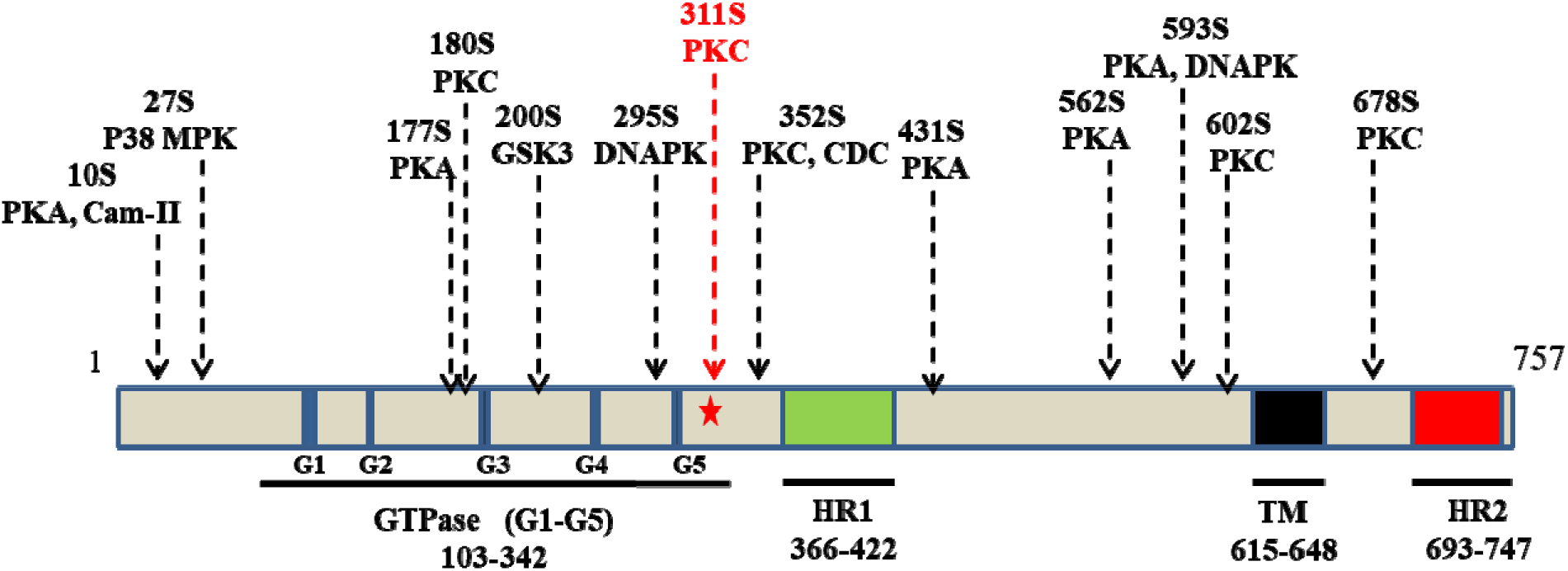
Diagrammatic representation of predicted serine phosphorylation sites and respective phosphokinases in the mutated MFN2 protein. The N311S mutation is shown with an asterisk. The prediction was performed by the NetPhos3.1 server.

### 3.5 Impact of N311S variant on the expression and localization of MFN2 protein

The expression of WT and MFN2-MUT proteins by stably transfected in H9c2 cells was evaluated by Western blotting, which showed significant differences in the level of protein expression. The level of MFN2 protein was down-regulated by 1.83 fold (p<0.003) due to the variants p.N311S as compared to WT (Figure 6A-B), which is a 45% reduction in the expression level compared to wild type.

**Figure 6.**
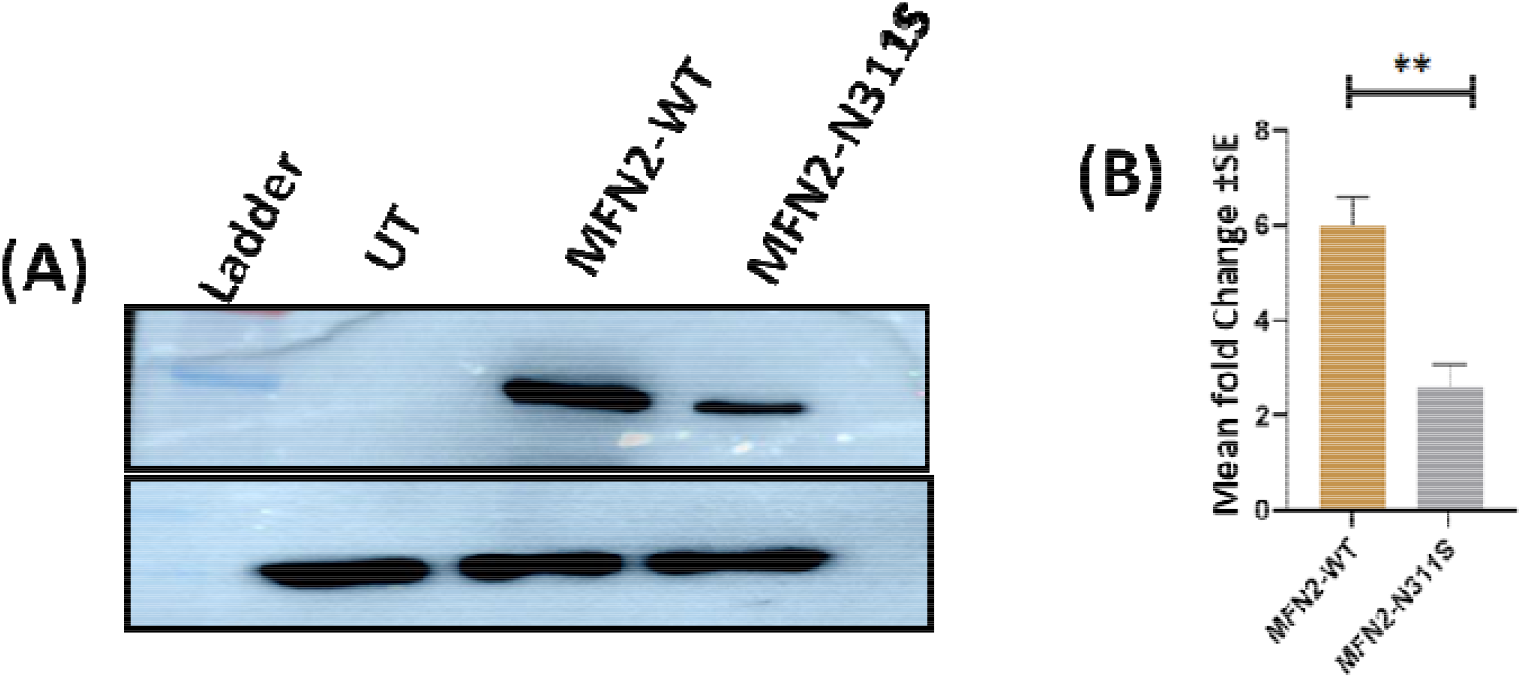
**(A)** Western blot analysis of wild-type and mutant protein showing expression of MFN2. **(B)** Representation of protein expression in terms of fold changes, calculated for each of the variant (P<0.05).

The cellular localization of MFN2 was studied by immunostaining which demonstrated mitochondrial localization of the protein in WT as well as in the MFN2-MUT (p.N311S) however, there was a reduction in the level of expression of protein in N311S as compared to WT.

Mito-Tracker staining demonstrated that MFN2-MUT cells exhibited a markedly altered clustered and clumped mitochondria. In MFN2-MUT showed perinuclear mitochondria accumulation in 58 ± 4% of cells, while fragmented mitochondria in 27 ± 6.2% of cells and reticulate in 18 ±3.4% of cells (Figure 7(A–B)). In contrast, WT cells primarily contained elongated tubular 76± 3.8%, fragmented 16±4.1% and clustered 10±3.4. TEM analysis demonstrated that MFN2-WT cells maintained elongated mitochondria with preserved ultrastructural organization, whereas MFN2-p.N311S mutant cells exhibited pronounced mitochondrial abnormalities, including fragmented and clustered mitochondria (Figure 8), suggesting impaired mitochondrial fusion and compromised mitochondrial architecture.

**Figure 7.**
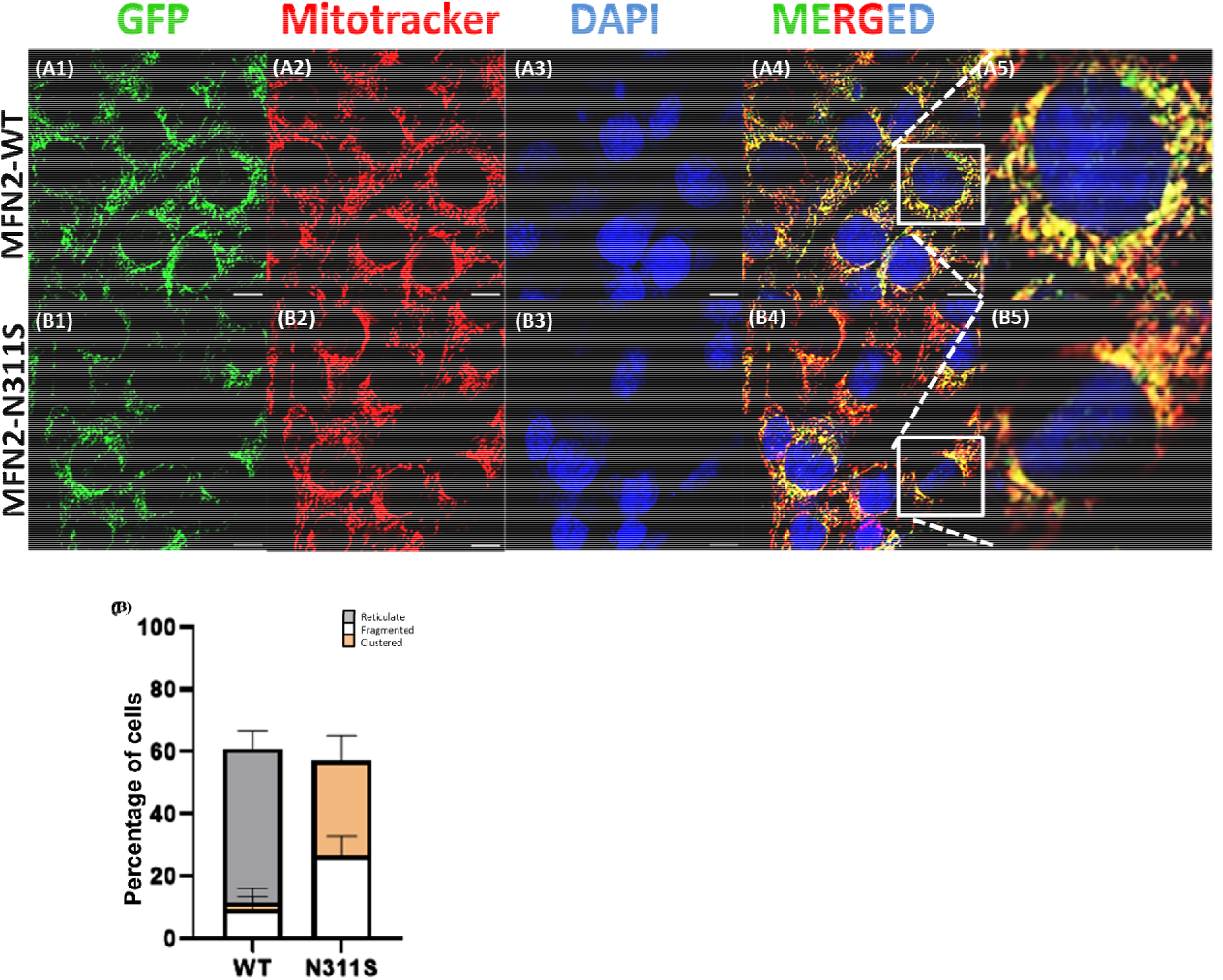
(**A)** Expression analysis of MFN2 protein (A1-B4) Immunostaining in H9C2 cells showing cellular localization of MFN2-WT and MFN2-MUT, A5 & B5 show higher magnification from boxed area. (**B)** Quantification of mitochondria morphology in the cell lines described, error bar represents mean ±SEM from n =3 separate blind experiments (∼100 cells per experiment).

**Figure 8.**
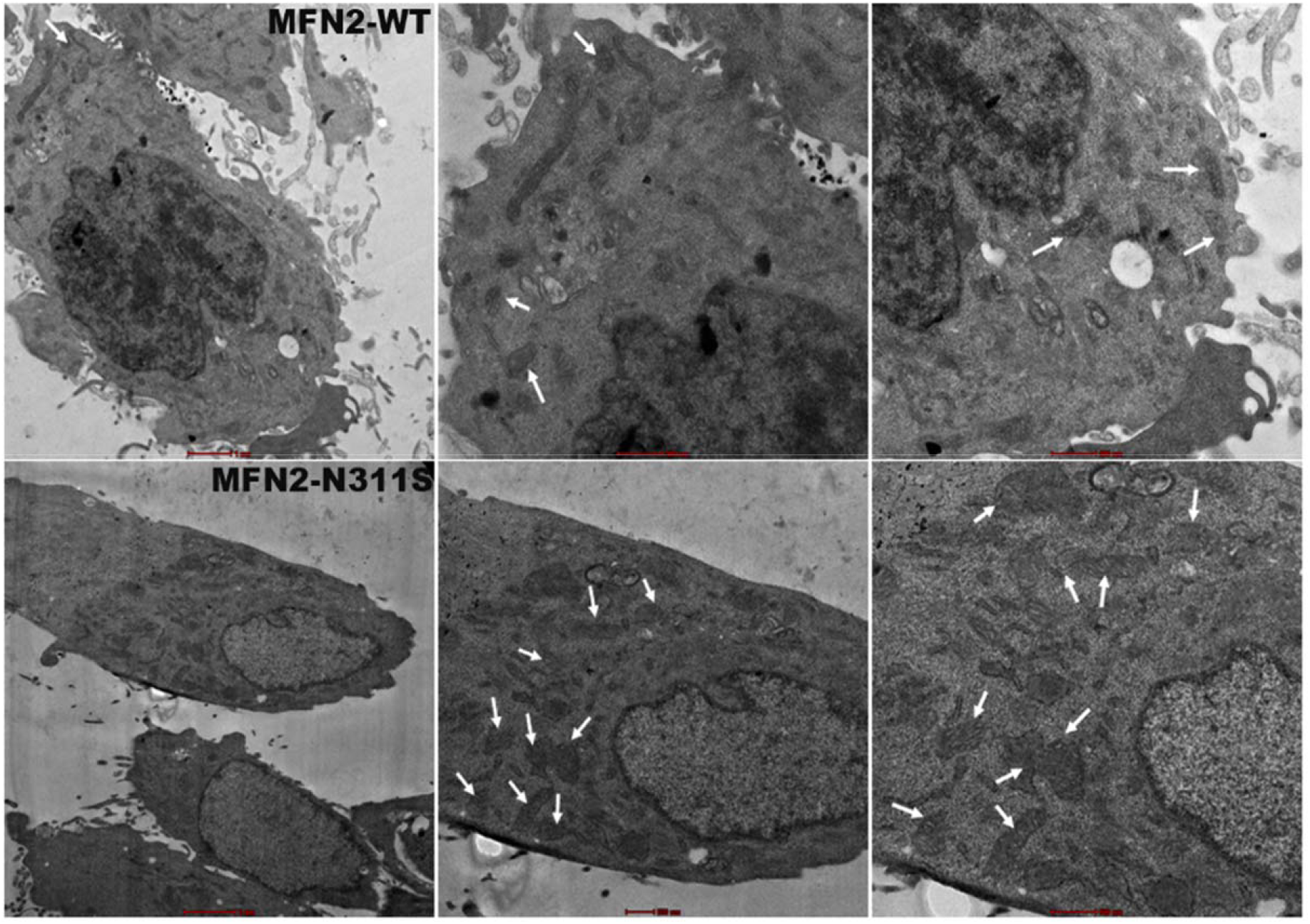
TEM images of stably transfected H9c2 cells with MFN2-WT and MFN2-N311S cells. White arrows correspond to mitochondria.

### 3.6 Impact of the MFN2 N311S Variant on Mitochondrial Mass, Bioenergetics, and Cell Survival

An MTT-based cell viability experiment was conducted to assess the effect of the N311S variant in the MFN2 gene on cellular health, utilising stably transfected MFN2-WT and MFN2-MUT (N311S) H9C2 cell lines (Figure 9A). No notable variation in cell viability was seen between WT and MUT cells, indicating that the mutation does not have an immediate cytotoxic impact.

**Figure 9.**
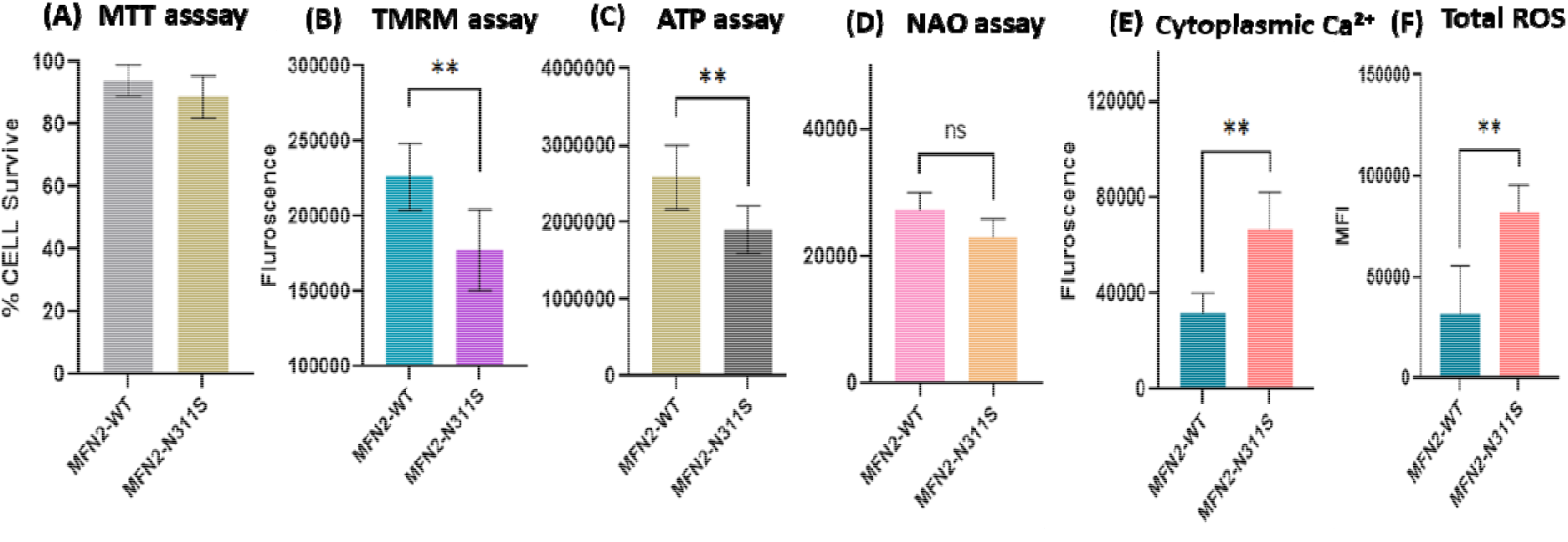
Effect of N311S on cell viability, mitochondria membrane potential, ATP level, mitochondrial mass, cytoplasmic calcium level and total ROS. **(A)** Analysis of cell viability using MTT assay **(B)** Analysis of mitochondrial membrane potential: histogram shows fluorescent intensity values **(C)** Luciferase assay to measure ATP level: histogram shows bioluminescence intensity-based-total ATP level **(D)** Mitochondrial mass assay **(E)** Cytoplasmic Ca^+2^ level measurement between MFN2-WT and N311S **(F)** Measurement of total ROS level. Mean values and standard deviations were calculated from at least 6 independent experiments., *p < 0.05. **p<0.01 .

To evaluate mitochondrial integrity, mitochondrial membrane potential (ΔΨm) was quantified using TMRE (tetra-methyl-rhodamine methyl ester), a fluorescent dye that accumulates in mitochondria in a manner dependent on membrane potential. A significant decrease in ΔΨm wa noted in MFN2-MUT (N311S) cells relative to WT cells (p < 0.01) (Figure 9B), signifying mitochondrial depolarisation. The loss of membrane potential is recognised to hinder oxidative phosphorylation and ATP synthesis.

Oxygen consumption rate (OCR) was assessed by a mitochondrial stress test assay to evaluate the impact on mitochondrial bioenergetics. Mitochondrial respiration was evaluated with the sequential administration of oligomycin (an ATP synthase inhibitor), FCCP (a protonophore), and the electron transport chain inhibitors rotenone and antimycin A. Respiratory metrics, including routine respiration, ATP-linked respiration (OXPHOS), leak respiration, and residual oxygen consumption (ROX), were assessed. High-resolution respirometry analysis revealed a marked decline in mitochondrial respiratory function in MFN2-MUT cells relative to MFN2-WT controls. Routine respiration was markedly reduced (p < 0.05), signifying impaired basal mitochondrial function. ATP-linked respiration (OXPHOS) was significantly decreased (p < 0.01), indicating compromised oxidative phosphorylation and lower ATP production capability. Leak respiration was markedly diminished in mutant cells (p < 0.05), possibly indicating modified proton conductance or decreased substrate oxidation. Moreover, residual oxygen consumption (ROX) was markedly reduced in MFN2-MUT cells (p < 0.01) (Figure 10A-C). Collectively, these results demonstrate a significant disruption in mitochondrial bioenergetics attributable to the N311S mutation.

**Figure 10.**
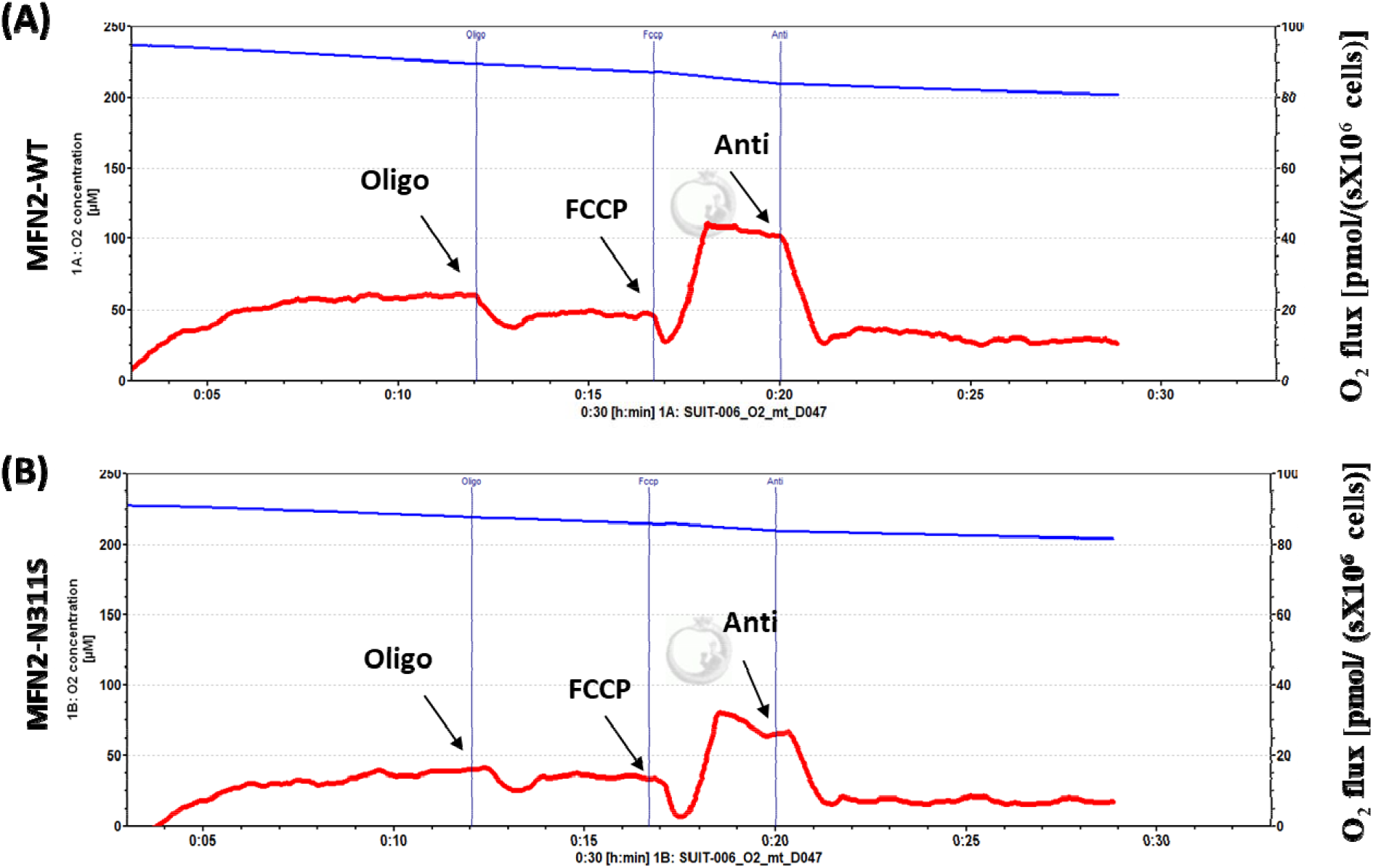
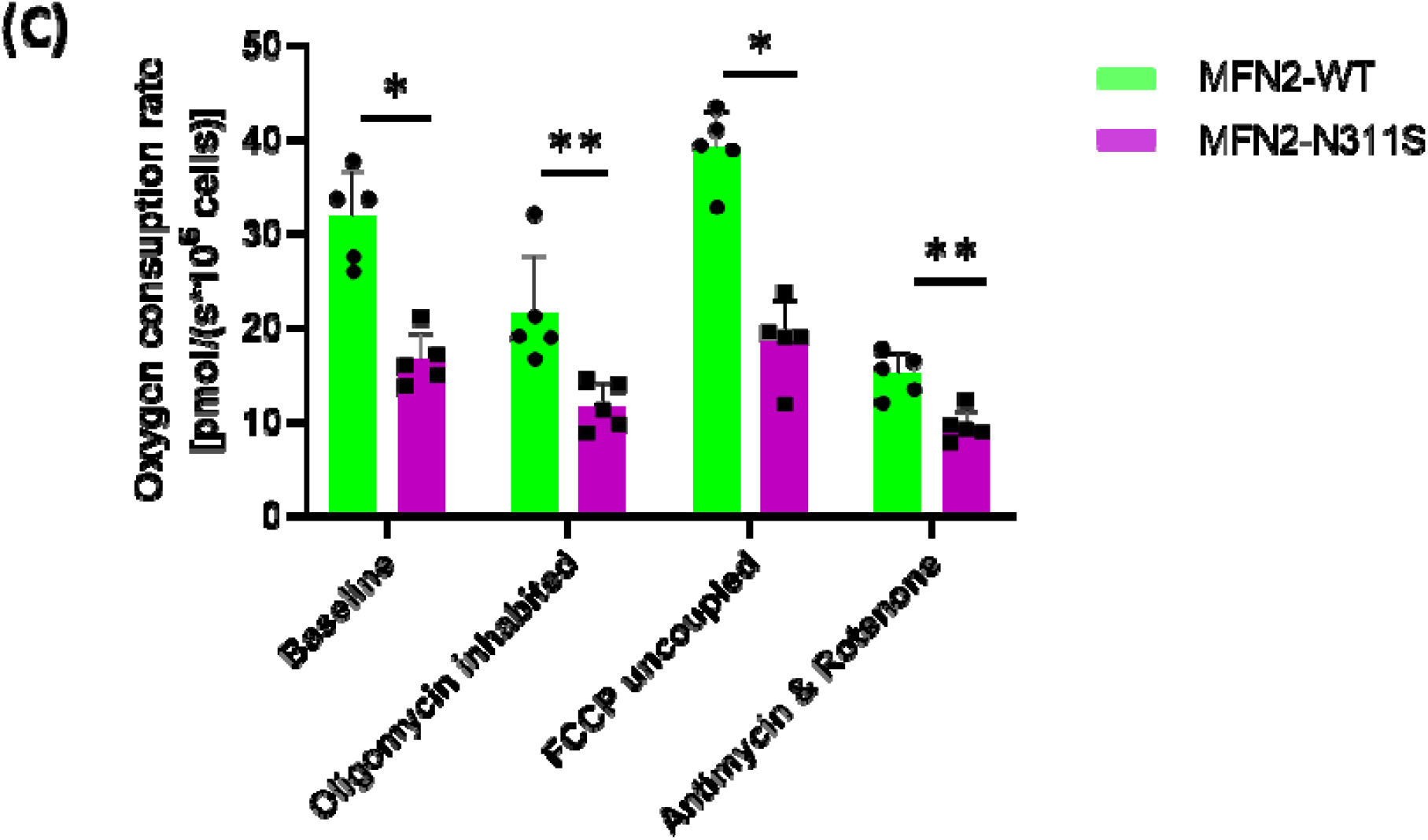
**(A & B)** Representative polarograms showing oxygen flux or oxygen concentration in H9C2 cells suspended in respiration buffer. Stable cell lines expressing MFN2-WT and MFN2-MUT were introduced into the oxygraph chamber. The blue tracing represents oxygen concentration within the chamber while the red trace indicates the rate of oxygen consumption (oxygen flux) by the cells. **(C)** Corresponding bar graph showing routine respiration, ATP-linked respiration (OXPHOS), leak respiration (LEAK) and residual oxygen consumption (ROX) rates in MFN2-WT and MFN2-N311S cells. Data are presented as mean ± standard error of mean. Significance in difference of means was tested by using two-way ANOVA followed by Tukey’s multiple comparison test. *p < 0.05. **p<0.01 . (WT = wild-type). Anti: antimycin; Oligo: oligomycin.

In accordance with the noted decline in OXPHOS, intracellular ATP concentrations were significantly diminished in MFN2-MUT (N311S) cells (p < 0.01), so further substantiating impaired mitochondrial energy synthesis. The association between the reported functional deficits and changed mitochondrial abundance was evaluated by assessing mitochondrial mass using 10-N-nonyl acridine orange (NAO), a cardiolipin-binding fluorescent dye that labels mitochondria irrespective of membrane potential (Maftah et al., 1989). Marginal reduction in mitochondrial mass was seen MFN2-MUT cells compared to MFN2-WT (Figure 9C-D).

### 3.7 Calcium homeostasis and ROS generation

With a hypothesis that pathogenic mutations in MFN2 could affect MAM-mediated Ca^2+^trafficking, we first loaded cells with Fura-2-AM and measured cytosolic Ca^2+^, which revealed that there is significant increase in cytosolic Ca^2+^ (p<0.01) as well as total ROS(p<0.01) content in MFN2-MUTas compare to MFN2-WT (Figure 9E-F).

### 3.8 Effect of MFN2 variant on Pi3K/AKT signalling and Hypertrophic gene expression

Mitofusin2 act as negative regulate of Pi3K/AKT signalling pathway (Guo et al., 2007; Fang et al., 2016). Shen et al., (2007) found that elevated level of Mfn2 expression promotes apoptosis via suppression of Akt activation in cultured cardiac myocytes cells. In order to verify this, we have checked the expression of Pi3k, Akt1and mTOR, which revealed a significant increase in expression of Pi3k (1.36-fold change, p<0.01), Akt1 (1.77-fold change p<0.01), mTOR (1.50-fold change, p<0.01). Similarly, Bax/Bcl2 ratio also showed insignificant change in N311S mutant as compare to MFN2 wild type (Figure 11). We further checked the expression level of hypertropic marker genes, which also showed marked elevation in i.e., Myh6(1.61-fold change, p<0.01), Nappa(1.43-fold change, p<0.05), Nfactc1(1.55-fold change, p<0.01) and Nfactc2(1.55-fold change, p<0.01), no significant change was observed in Actc1 in N311S mutant as compare to MFN2 wild type (Figure 12).

**Figure 11.**
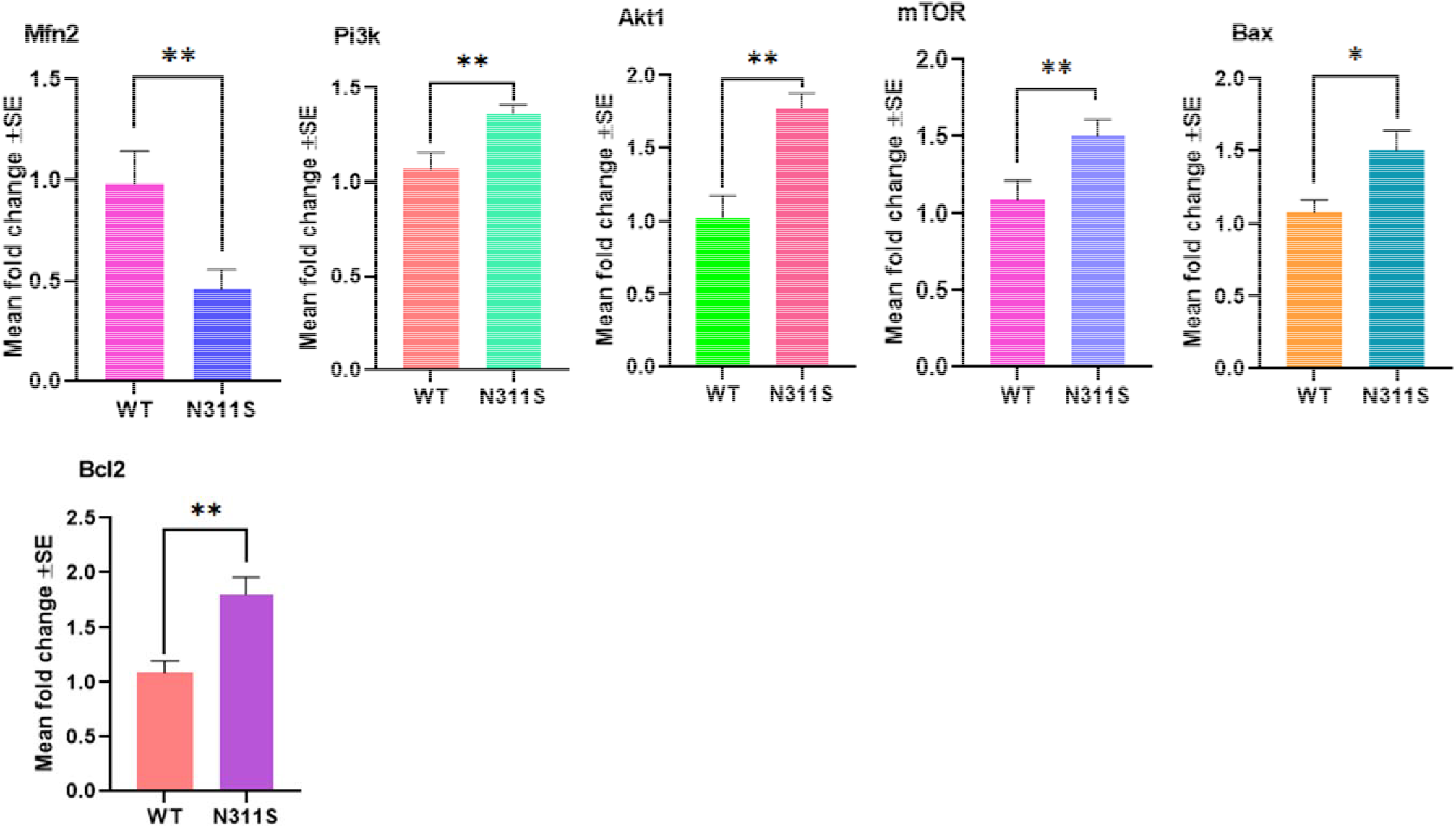
qPCR assay showing expression of target genes Pi3k, Akt1, mTOR, Bax and Bcl2 in response to MFN2 wild-type and N311S mutant protein. Results shown are mean fold change ± SE of mean for three independent experiments performed in triplicates, *p < 0.05. **p<0.01.

**Figure 12.**
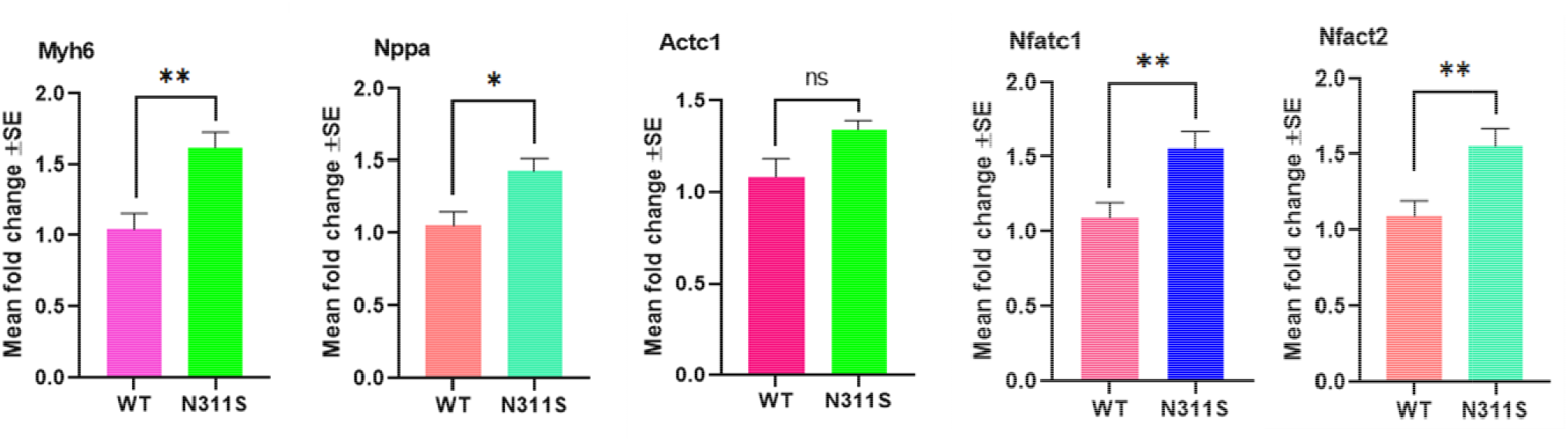
qPCR assay showing expression of cardiac-enriched target genes Myh6, Nppa, Actc1, Nfact1 and Nfact in response to MFN2 wild-type and N311S mutant protein. Results shown are mean fold change ± SE of mean for three independent experiments performed in triplicates, *p < 0.05. **p<0.01.

### 4.0 Discussion

*MFN2* (HGNC: <u>16877</u>) is a nuclear gene, encoding a mitochondrial outer membrane anchoring protein, localized to chromosome 1p36.22. The protein MFN2 along with its paralog, MFN1 operate as membrane bound GTPases, mediating mitochondrial fusion and clustering. Apart from mitochondrial fusion, MFN2 also involved in tethering of mitochondria–ER (Brito & Scorrano, 2008), calcium homeostasis, neuroaxonal transport (Misko et al., 2010), mitophagy, cellular apoptosis and cell proliferation (Chen et al., 2014).

In the present study, we have identified a rare missense variation c.932A>G; p.(N311S) through WES, in a patient with idiopathic DCM, confined to the GTPase domain of MFN2. This variation is absent in 100 ethnic-matched heathy control from the same geographical location as well as in databases namely 1000 genome and Genome Asia 100K, while present at an extremely low allele frequency in gnomAD (0.00000812), and absent in Indian databases (Indigenomes, INDEX-db). Multiple bioinformatic tools applying ‘VarCard’ analyses, have predicted deleterious nature of this variant. Further, using ‘Protscale’ tool, significant changes in the hydrophobicity, total beta strand and alpha helix are observed in MFN2 mutant protein when compared to wild type. Prior research has confirmed the pathogenic significance of *MFN2* mutations, which are known to be frequently associated with several neuromuscular diseases namely Charcot–Marie–Tooth disease (Züchner et al., 2004; Verhoeven, 2006; Franco et al., 2022), Parkinsons disease (Celardo et al., 2016), Alzheimer’s disease,(Wang et al., 2009) and optic degeneration (Züchner et al., 2006). Besides these, MFN2 mutations have also been linked to cardiomyopathy together with mitochondrial dysfunction (Franco et al., 2023). A human MFN2 mutation, p.R400Q, in the HR1 domain induces cardiomyopathy marked by mitochondrial depolarisation and reduced mitochondrial respiration, without affecting mitochondrial motility. The introduction of the R400Q mutation into the mouse *Mfn2* gene using CRISPR editing, lead to neonatal cardiomyopathy characterised by cardiac-restricted disease. Moreover, the expression of the identical mutation in a Drosophila model (R400Q) resulted in significant mitochondrial fragmentation, heart dilatation, diminished fractional shortening, and compromised locomotor activity (Eschenbacher et al., 2012). The conditional double KO of Mfn1 and Mfn2 also exhibited DCM phenotype in mouse heart (Chen et al., 2011).

The variant identified in this study, c.932A>G; p.(N311S), is localised to GTPase domain. Mutations in the highly conserved GTPase domain of MFN2 hinder mitochondrial fusion and subsequently compromise mitochondrial function (Züchner et al., 2004; Detmer & Chan, 2007). While the GTPase domain is crucial for MFN2-mediated mitochondrial fusion and membrane tethering (Detmer & Chan, 2007; Tilokani et al., 2018) and nearly 100 mutations in this domain are associated with human diseases, particularly in CMTA2. Only a limited number of disease-associated MFN2 mutations have undergone functional (molecular/ biochemical) characterization. The variant p.(N311S), in the present study, has shown a significant reduction in expression of mutant protein (1.83 fold) as well as in MFN2-MUT transcript (∼ 2.15 fold; qRT-PCR). Binding affinity of MFN1 is also predicted to be decreased with MFN2-MUT compared to WT (Table 3). Furthermore, a significant functional impairment is also observed in mitochondrial integrity (ΔΨm), and intracellular ATP concentrations, while a modest decrease in mitochondrial mass in mutant MFN2. N311S-expressing cells demonstrated a pronounced decrease in TMRE intensity, signifying substantial mitochondrial depolarisation and compromised mitochondrial coupling efficiency. In accordance with the reduction of ΔΨm, intracellular ATP concentrations were markedly diminished in MFN2-MUT cells relative to MFN2-WT controls, indicating impaired bioenergetic performance. Comprehensive evaluation of mitochondrial respiratory function using oxygen consumption rate (OCR) analysis further substantiated these defects. Mitochondrial stress test estimating oxygen consumption rate (OCR), has demonstrated significant depletion in basal respiration, ATP-coupled respiration, and lowed residual oxygen consumption (ROX) in cells expressing mutant MFN2-N311S. The decreased ATP-linked oxygen consumption rate portion specifically signifies compromised oxidative phosphorylation efficiency, while the decline in spare respiratory capacity implies a restricted capacity of mutant cells to meet heightened energy demands. All these observations are corroborated by *in silico* protein stability predictions via DYNAMUT2, MUPro, DUET, I-Mutant, CUPSTAT, SDM, which suggested that the N311S change reduces protein stability. Moreover, the superimposed 3D structures of MFN2 N311S mutant versus wild-type show pronounced conformational changes between the two models, as reflected by a root mean square deviation (RMSD) value of 8.951 Å, indicative of substantial atomic displacement. Such a high RMSD value suggests that the mutation induces remarkable conformational alterations, potentially disrupting the native folding pattern, domain orientation, and interaction interface of MFN2, which may in turn compromise its functional association with partner proteins. Overall, the reduced protein expression and the associated functional deficits are likely to arise from structural destabilisation of the mutant protein . These data collectively indicate that the N311S mutation significantly undermines mitochondrial bioenergetic integrity, resulting in compromised oxidative phosphorylation, decreased coupling efficiency, and lower cellular energy stores.

**Table1.** In-silico analysis of the identified *MFN2* variant across different population database.

| Nucleotide change | Amino Acid change | dbSNP ID | Type of mutation | MFN2 Domain | Status | ClinVar Annotaion | HGMD_Disease | Indigenomes | 1000 Genomes | Genome Asia 100K | gnomAD |
| --- | --- | --- | --- | --- | --- | --- | --- | --- | --- | --- | --- |
| c.932A>G | p.Asp311Ser | rs748838916 | Non-Synonyms | GTPase | Known | NR | NR | NR | NR | NR | 0.00000812249 |

Abbreviations: dbSNP: Single nucleotide polymorphism database, NR: Not reported; HGMD: Human Gene Mutation Database;

**Table2.** Prediction of pathogenic likelihood of *MFN2* variant using VarCards and structure stability assessment.

| Variant | SIFT | PolyPhen2 | LFT pred | Mutation tester | FATHMM | PROVEN | CADD | DYNAMUT2 | MUpro | CUPSAT | DUET | I-Mutant | SDM | mCSM |
| --- | --- | --- | --- | --- | --- | --- | --- | --- | --- | --- | --- | --- | --- | --- |
| c.932A>G | Damaging | Damaging | Damaging | Disease causing | Damaging | Damaging | Damaging | -0.89 kcal/mol<br>Destablizing | -0.89 kcal/mol<br>Destablizing | -0.49 kcal/mol<br>Destablizing | -0.995 kcal/mol<br>Destablizing | Decrease | -0.85 kcal/mol<br>Destablizing | -1.0.58 kcal/mol<br>Destablizing |

Abbreviations: SIFT: sorting intolerant from tolerant; PolyPhen2: polymorphism phenotyping v2; FATHMM: functional analysis through hidden Markov models; CADD: combined annotation dependent depletion Dynamut2: dynamic Mutant 2; MUpro: mutant protein predictor; CUPSAT: Cologne University Protein Stability Analysis Tool; DUET: Double Mutation Estimator Tool; I-mutant: internet-based mutant; SDM: site-directed mutator; mCSM: mutation cutoff scanning matrix.

**Table3.** Interaction of MFN1 with MFN2-WT and MFN2-MUT.

| Parameter | MFN2-WT - MFN1 | MFN2-N311S - MFN1 |
| --- | --- | --- |
| Interface area (Å<sup>2</sup>) | 958.4 | 940.4 |
| ΔG (kcal/mol) | -9.2 | -8.8 |
| ΔG P-value | 0.371 | 0.369 |
| Hydrogen bonds (NHB) | 6 | 7 |
| Salt bridges (NSB) | 3 | 5 |
| Interface residues (MFN2) | 28 | 29 |
| Interface residues (MFN1) | 31 | 29 |
| Solvation free-energy gain (MFN2) | -3.6 kcal/mol | -4.0 kcal/mol |
| Solvation free-energy gain (MFN1) | -5.6 kcal/mol | -4.3 kcal/mol |

In addition to bioenergetic deficiencies, in this study significantly higher level of ROS and Ca+2 is observed in case of MFN2-MUT (N311S). The mutant cells exhibited heightened cytosolic Ca² concentrations and augmented reactive oxygen species (ROS) generation. Mitochondria are pivotal in cytosolic Ca² buffering via interactions with the endoplasmic reticulum (ER) at mitochondria-associated membranes (MAMs) (Patergnani et al., 2011; Rizzuto et al., 2012; Filadi et al.,2015). Essential elements in this process comprise the inositol 1,4,5-trisphosphate receptor (IP3R) located on the SR/ER membrane, the voltage-dependent anion channel (VDAC) situated on the outer mitochondrial membrane (OMM), and the mitochondrial calcium uniporter (MCU) found on the inner mitochondrial membrane (IMM) (Inagaki et al., 2022). MFN2 ablation in mouse embryonic fibroblasts and HeLa cells augmented ER–mitochondria separation, leading to increased cytosolic Ca², and heightened ROS generation (Brito and Scorrano 2008; Cosson et al. 2012; Ngoh, et al. 2012; Filadi et al. 2015). These findings corroborate our observation that the N311S mutation modify ER-mitochondria tethering, enhance cytosolic Ca² and elevate oxidative stress. Increased cytosolic Ca² can diminish mitochondrial metabolic efficiency and enhance ROS production, thereby undermining oxidative phosphorylation. Mitochondrial Ca² mechanistically modulates tricarboxylic acid cycle dehydrogenases and enhances electron transport chain activity and ATP generation. Consequently, dysregulated Ca² handling in MFN2-MUT cells likely leads to the observed reduction in ΔΨm, ATP generation, and ATP-linked respiration. The findings indicate that the N311S mutation interferes with MFN2-mediated communication between the endoplasmic reticulum and mitochondria, leading to calcium imbalance, oxidative stress, and compromised mitochondrial bioenergetics. Deficiency of Mfn2 in knockout mouse model, implicated generation of ROS as a consequence of adoptive mitochondrial respiration in stressful condition leading to mitophagy or apoptosis (Song et al., 2014). Similarly, cellular stress generating excessive ROS is known to induced Ca² release into the cytoplasm either from extra cellular compartment or from Ca² storage organelle, the ER/SR. Excessive extracellular Ca², further accelerate ROS production, thus inducing a vicious cycle of ROS generation and Ca² overload which possibly lead to disease pathogenesis.

Decreased MFN2 expression is recognised to hinder mitochondrial fusion and enhance mitophagy, aligning with its critical function in preserving mitochondrial network integrity and quality control (Chen & Dorn, 2013; Song et al., 2014).The impaired MFN2 function leads to fragmented mitochondria, while its overexpression facilitates the development of a hyperfused mitochondrial network (Koshiba et al., 2004). The clustered and clumped mitochondria observed in this study as consequence of MFN2-MUT (N311S) compared to WT, clearly implicate mitochondrial fragmentation. Further, MFN2-MUT showed perinuclear mitochondria accumulation (Figure 8). Drosophila models harbouring MFN2 mutations, specifically R94Q and T105M, demonstrate aggregation of unfused mitochondria and altered network architecture. Electron microscopy examinations of nerve samples from CMT2A patients have indicated the presence of localised mitochondrial aggregation, hence reinforcing the pathogenic significance of mitochondrial clustering (Verhoeven, 2006; Fissi et al., 2018). Reduced mitochondrial Mfn2, causing fragmented and, clustered mitochondria further increasing ER-mitochondrial coupling, supported by the perinuclear localisation. Further, ER-mitochondrial coupling, increase Ca² transfer to mitochondria, causing Ca² overload and cell death (Filadi et al., 2015).

Herein, the putative phosphorylation of the Ser residue at aa position 311 originated due to p.N311S substitution, is predicted to be phosphorylated by PKC and other unspecified kinases with prediction score of 0.616 and 0.871 respectively. Evidences from the literature have shown that phosphorylation of MFN2 mediated by PINK1, kinase promote MFN2-PARKIN binding (Chen & Dorn, 2013). Phosphorylation of MFN2 T111, S378 and S442 has been shown to impede mitochondrial fusion, compromising oxidative phosphorylation, ATP generation and mitochondrial depolarization leading to mitophagy by PARKIN-mediated ubiquitination stimulating proteasomal degradation (Detmer & Chan, 2007; Li et al., 2022). Our *in silico* docking data indicated a significant disparity in contact stability between the wild-type and mutant MFN2 proteins. Further molecular docking analyses between MFN2 and PRKN revealed a distinct difference in the stability of their interactions between the wild-type (WT) and mutant (MUT) forms of MFN2. The MFN2-WT complex exhibited a greater number of hydrogen bonds and a more robust interfacial network, indicating enhanced binding affinity and structural stability compared to the MFN2-MUT complex. Intriguingly, the mutation induced a spatial reorganization of the interaction interface, with a complete shift of the binding residues from the N-terminal GTPase domain in the wild-type to the C-terminal heptad repeat (HR) domains in the mutant protein. This domain-level transition suggests that the mutation may alter the conformational landscape of MFN2, thereby affecting its ability to engage effectively with PRKN. Defective PRKN recruitment is likely compromise PRKN mediated mitophagy.

Besides its well-established role in mitochondrial fusion and organelle communication, MFN2 has also been reported to modulate the PI3K/AKT signalling axis. In particular, MFN2 can suppress mTORC2-dependent AKT activation through interaction with Rictor, suggesting a context-dependent inhibitory influence on AKT signalling (Xin et al., 2021; Xu et al., 2017) In accordance with diminished MFN2 expression in mutant N311S cells, we noted a substantial elevation of PI3K (1.36-fold), AKT1 (1.77-fold), and mTOR (1.50-fold), indicating transcriptional activation of the PI3K/AKT/mTOR pathway. Notwithstanding this seemingly pro-survival signalling, apoptotic markers Caspase-3 (1.38-fold) and Caspase-9 (1.67-fold) were dramatically increased, whereas the Bax/Bcl-2 ratio exhibited no change. This may indicate activation of the intrinsic apoptotic pathway due to mitochondrial stress, potentially caused by mitochondrial depolarisation, reactive oxygen species buildup, and calcium dysregulation. Consequently, compensatory survival signalling may be inadequate to mitigate mitochondria-induced apoptotic stress.(Green & Reed, 1998; Manning & Toker, 2017).

In light of the strong association between mitochondrial dysfunction and cardiac remodelling, we conducted a thorough assessment of hypertrophic gene expression (Dorn, 2015). MFN2-MUT cells demonstrated substantial overexpression of *Myh6, Nppa, Nfatc1*, and *Nfatc2,* whereas *Actc1* expression remained constant. The *Nppa* and *Myh6* serve as traditional indicators of cardiac remodelling, whilst NFAT transcription factors facilitate Ca² -dependent hypertrophic signalling. Increased cytosolic Ca² concentrations in N311S cells may consequently initiate NFAT-mediated transcriptional pathways, connecting mitochondrial Ca² dysregulation to hypertrophic gene expression. Furthermore, PINK1-PRKN-MFN2 mediated mitophagy is likely associated with cardiac remodelling leading to cardiomyopathy.

## Conclusion

Collectively, our results demonstrate that the N311S variation diminishes MFN2 protein stability, hinders mitochondrial fusion, affects ER–mitochondria Ca² signalling, and undermines oxidative phosphorylation. These modifications are associated with oxidative stress, apoptotic signalling, activation of components within the PI3K/AKT/mTOR pathway, and the promotion of hypertrophic gene expression. This combined bioenergetic and signalling imbalance may render cardiomyocytes susceptible to pathological remodelling. Given that modified MFN2 expression and function are associated with cardiovascular conditions like cardiac hypertrophy, atherosclerosis, and pulmonary arterial hypertension. Our research highlights the essential role of MFN2 in preserving mitochondrial integrity, calcium homeostasis, and cellular energy balance.

## Data Availability

All data produced in the present study are contained within the manuscript and its supplementary materials.

## Acknowledgment

We sincerely thank the patients and their families for their invaluable participation in this study. We are grateful to Padma Shri Prof. T. K. Lahiri and Prof. D. Agrawal, Department of Cardiothoracic and Vascular Surgery, for their constant encouragement and support in patient enrolment. We also acknowledge Prof. Pozzan’s Laboratory, University of Padova, Italy, for providing the full-length MFN2 clone. We thank the Sophisticated Analytical Instrument Facility (SAIF), AIIMS, New Delhi, for TEM facilities, and the Sophisticated Analytical & Technical Help Institute (SATHI), Banaras Hindu University, Varanasi, for access to the laser-scanning super-resolution microscopy system. We are grateful to Prof. D. Das, Department of Biochemistry, Institute of Medical Sciences, Banaras Hindu University, Varanasi, for providing access to the Oxygraph-2k system (Oroboros Instruments, Innsbruck, Austria). This work was supported by the Department of Biotechnology (DBT), Government of India (BT/PR12369/MED/12/678/2014), and by an ICMR Senior Research Fellowship awarded to Ms. Mohini Gupta (F. No. 2021-11036/Proteomics-BMS).

## Author contributions

Bhagyalaxmi Mohapatra: Conceptualization, planning and supervision, Experimental design, validation, resources, Preparation, reviewing & editing of manuscript; Mohini Gupta: Data curation, execution of experiments, validation, software handling, primary draft preparation of manuscript. Amrita Mukhopadhyay: Input in experimental design, 3-D Modelling and alignments. Manohar Lal Yadav: NGS data analysis. Dharmendra Jain: for identifying and enrolling patients for the study.

